# Association between driving status and visiting places among older adults in a suburban area in Japan: Findings from a cross-sectional survey

**DOI:** 10.1101/2024.08.20.24311843

**Authors:** Taiji Noguchi, Ayane Komatsu, Sayaka Okahashi, Takeshi Nakagawa, Xueying Jin, Yumi Shindo, Tami Saito

**Author notes:** **Corresponding author:** Tami Saito, PhD, Department of Social Science, Center for Gerontology and Social Science, Research Institute, National Center for Geriatrics and Gerontology, 7-430 Morioka, Obu, Aich 474-8511, Japan. These authors contributed equally to the work (Taiji Noguchi, Ayane Komatsu).

## Abstract

**Introduction:** Driving is an important mobility resource of increased outings and social activities among older adults; yet, little is known about the impact of driving restrictions on visiting places. We examined the association between driving status and the number of visiting places and the moderating role of alternative transportation use.

**Methods:** This cross-sectional study recruited community-dwelling individuals aged 65 years and above with functional independence from a suburban area through a mailed self-administered questionnaire. Visiting places were scored by assessing a total of 34 specific places over the past year using the Participation in ACTivities and Places OUTside Home Questionnaire (ACT-OUT); four subdomains of these paces were also measured: (A) consumer, administration, and self-care places (e.g., grocery shop, hairdresser, bank, post office, and government office); (B) places for medical and health care (e.g., the dentist or doctor’s office and hospital); (C) social, cultural, and spiritual places (e.g., family, relative and friend’s home, restaurant, cafe, and entertainment and cultural places); and (D) places of recreation and physical activities (e.g., park, sports facility, and forest, mountain, and sea). Driving status (self-driving or not) and other available transportation options (public transportation and ride-sharing with family/friends) were assessed.

**Results:** Data from 432 individuals were analyzed (mean age 74.8 years; 52.8% women). Multivariable linear regression analysis revealed that not driving was associated with lower scores of visiting places (β=-0.40, *P*<0.001). For subdomains, not driving was associated with lower scores for consumer, administration, and self-care places (β=-0.32, *P*=0.007) and social, cultural, and spiritual places (β=-0.44, *P*<0.001). Sensitivity analysis with inverse probability weighting confirmed the robustness of these results. Public transportation availability (trains and buses) moderated the association between not driving and visiting places.

**Conclusions:** These findings suggest that driving restrictions have the potential to reduce the visiting places among older adults, particularly life-related and social and cultural places.

## Introduction

As population aging globally proceeds, car driving among older adults gathers more attention. Older drivers may face an increased risk of fatal car accidents than younger drivers [1] and experience more care accidents as their physical and cognitive functions decrease [2]. Conversely, driving cessation in older adults raises several concerns: the impact of driving cessation on their health and well-being and alternative transportation after driving cessation to maintain their mobility [3, 4].

Driving generally ameliorates their physical and mental health [5]. This is in contrast with observations among younger drivers, who are sometimes at higher risk of a sedentary lifestyle or obesity [6]. For at least partial older adults, driving means autonomy and identity [7] and enables them to continue their independent living in the communities, particularly when they live in suburban and rural areas [8, 9]. Indeed, several longitudinal studies show that older drivers demonstrate lower risks of functional decline than non-drivers [10, 11]. Additionally, older drivers are less likely to have social risk factors for functional decline, such as restrictions on life-space mobility [12-15], social networks [16], companionship with friends [15], and social activities [16, 17] than non-drivers. However, it remains unclear where older adults can or cannot continue to visit after they have stopped driving.

Studies have highlighted the importance of places for older adults. Those from environmental gerontology have delved into the meaning of places or aging-in-place and stated that to older adults, places do not merely mean geographical spaces but also involve affective, cognitive, behavioral, and social links that are formed from their environment-related experiences [18]. As people live in a place for a long time, they develop a strong sense of connection, such as attachment to places and residents living there [19]. Such a connection is also closely linked to place identity, a sub-identity of the self [20]. These emotional and cognitive links to places foster the motivation for independence and autonomy [18]. In a more behavioral sense, visiting places can increase engagement in meaningful activities that older adults perceive, such as self-care, mobility, domestic life, work/education, interpersonal interaction, social life, sports, and leisure [21]. However, places generally become less accessible when one stops driving, unless they are located within walking distance, or when alternative transportation is available. It has been shown that non-driving older adults are less likely to travel for leisure [22]. Assessing visiting places and the decrease after driving cessation has a potential as a mechanism between driving and functional health in older adults.

While places have multiple meanings for older adults, the places visited by older adults can also infer accessibility, particularly when older adults stop visiting due to a functional decline or restriction of mobility, such as driving cessation. The concept of age-friendly cities proposed by the World Health Organization (WHO) underscores the importance of convenient and accessible locations for social activities that cater to older adults’ needs and preferences [23]. Thalén et al. reported in their cross-national study that cognitive decline could impact out-of-home participation differently according to the purpose of the activities and accessibility of transportation [24]. Although the impacts of cognitive decline and driving cessation can be distinct, assessing visited places and their differences according to driving status can provide practical information for promoting more accessible communities.

Alternative transportation is another issue related to driving cessation among older adults. WHO also suggests in their guideline for age-friendly cities that accessible and affordable transportation systems should be equipped [23]. While both public transportation and shared rides with family or friends are the most frequent alternative transportation means for older adults [25], they differ regarding flexibility and sense of independence in using. Public transportation may be less flexible in time than help from family or friends, however, it rarely compromises the older adults’ sense of independence. Hirai et al. indicated that the risk of functional decline was higher in former drivers relying only on ride-sharing with family or friends than in those with independent mobility such as bicycle or public transportation [11]. However, findings are still inconsistent regarding the moderation effect of public transportation or ride-sharing [26-28]; this necessitates further studies, including the association with visiting places.

Therefore, this study aimed to elaborate on which places older adults visit and examined whether these places differ according to their driving status. The second aim was to examine how public transportation use or ride-sharing with family or friends moderates the association between driving status and visiting places. This study adds empirical evidence that the non-driving of older adults influences the places to visit, with the roles of alternative transportation options.

## Methods

### Participants and procedures

This cross-sectional study was conducted as part of the National Center for Geriatrics and Gerontology-Universal Communities (NCGG-UniCo) project, an ongoing study aimed at improving the social participation and well-being of all people, including individuals with dementia, by developing supportive and creative communities. This study involved community-dwelling older adults with functional independence who lived in Obu City, Aichi, Japan. Japan is one of the most aging countries, promoting barrier-free urban development with a focus on facilitating mobility for older adults and individuals with physical disabilities [29]. Obu City, the survey area for the present study, has suburban residential and industrial areas, exhibiting the features of both urban and rural zones; geographically, the city is located in the center of Japan and experiences a typical Japanese climate [30]. The population of the city is approximately 90,000 (38,000 households), with an aging rate of 22% [31]. The percentage of individuals holding a driver’s license was 83%, and the percentage of those aged ≥65 years was 66% [32].

We conducted a self-administered mailed questionnaire survey between December 2023 and January 2024 among older adults who did not receive public long-term care insurance benefits. Of the 20,228 individuals aged ≥65 years registered in the municipality as of November 1st, 2023, 16,958 without public long-term care insurance benefits were eligible. Among them, 1,000 individuals selected by random sampling were distributed the questionnaire, and 686 responded (response rate: 68.6%). Of the respondents, we excluded 227 individuals provided no or invalid consent to participate in the research and 29 individuals requiring care in daily living. Thus, a total of 432 older adults were included in the final analysis.

### Visiting places

To assess visiting places, the ACTivities and Places OUTside Home Questionnaire (ACT-OUT), modified in a form applicable to the Japanese context, was used [24, 33]. Participants were asked about 34 specific places, such as the restaurant and family’s house, with a binary answer regarding whether or not they had visited these places in the past year. A list of these items is presented in Supplementary Table 1. The evaluated visiting places comprised four subdomains: (A) consumer, administration, and self-care places (9 items, including small grocery shop, supermarket, pharmacy, hairdresser, salon, barbershop, bank, post office, and government office); (B) places for medical and health care (5 items, including alternative adjuvant therapy, dentist and doctor’s office, hospital, and senior daycare); (C) social, cultural, and spiritual places (12 items, including family/relative and friend’s places, restaurant, cafe, building for worship, cemetery, entertainment, cultural places, historic building, and stadium, theater); (D) places of recreation and physical activities (8 items, including garden, park, green area, sports facility, cottage, and forest, mountain, lake, sea). We calculated the total score by adding a point for each of the 34 places visited (range 0–34 points; higher scores indicated more active); additionally, four subdomain scores were calculated (domain A, range: 0–9 points; B, range: 0–5 points; C, range: 0–12 points; and D, range: 0–8 points).

### Driving status and alternative transport means

Participants indicated their means of transportation available for outings by selecting from seven available means. This study focused on three modes of transportation: car driving, public transportation, and ride-sharing with family/friends. Regarding the status of car driving, participants answered whether or not they drove a car or a motorbike by themselves (yes=driving; no=not driving). Likewise, public transportation (e.g., public trains or buses) and ride-sharing with family/friends were also dichotomized, respectively (yes=available; no=not available).

### Covariates

The covariates included age, gender, living arrangement, educational attainment, equivalent household income, number of illnesses, motor function, subjective cognitive function, mental health, instrumental activities of daily living (IADL) performance, and independent mobility difficulties. Age was considered a continuous variable. Living arrangement was dichotomized into living together and living alone. Educational attainment (years) was categorized as <10, 10–12, ≥13, and other. Equivalent household income was calculated by dividing the income of each household by the square root of the household size (family member number) and was categorized as low (<2.00 million Japanese Yen), middle (2.00–3.99 million), and high (≥4.00 million). The number of illnesses from a list of 17, including cancer, stroke, heart disease, diabetes, digestive disease, dementia, and mental disorders, were categorized as none, one, two, and three or more. Motor function was assessed using a five-point subscale of the Kihon checklist (KCL), which is widely used as a screening tool for frailty [34], and dichotomized as not impaired (<3 points) and impaired (≥3 points). Subjective cognitive function was assessed using a 3-point subscale of the KCL, and dichotomized as not impaired (0 points) and impaired (≥1 points). Mental health was assessed using the Japanese version of the World Health Organization-Five Well-being Index (WHO-5-J) and dichotomized by a 13-point cutoff: not decline (≥13 points) and decline (<13 points) [35]. IADL performance was assessed using a 5-point subscale of the Tokyo Metropolitan Institute of Gerontology Higher Competence Scale, and those with difficulty in at least one item were categorized as with difficulty and others as without difficulty [36]. Independent mobility difficulties were assessed using a subitem on the mobility in the Japanese version of the five-level version of the EuroQol five-dimensional questionnaire (EQ-5D-5L) [37], and dichotomized as no (no problems) and yes (slight problems to unable to walk about).

### Statistical analysis

First, the characteristics of the participants were described. Second, the prevalence was calculated for 34 visits places. Third, to examine the association between driving status and visiting places, we applied multivariable linear regression analysis and obtained regression coefficients (βs) and 95% confidence intervals (CIs). Two analytical models were used: a crude model and a model adjusted for all the covariates. Primary analysis was conducted on all scores of visiting places, and secondary analysis was conducted on each of the four subdomain scores. In the analysis, the scores for visiting places pf all outcomes were entered into the model after standardization (mean=0 and standard deviation [SD]=1). We also performed several sensitivity analyses to test the robustness of the results. First, in order to estimate the potential outcomes after conditioning on covariates, an inverse probability weighting (IPW) model was adopted. We calculated the propensity scores for driving status using logistic regression analysis and performed the regression analyses with the IPW. Second, we conducted analyses restricted to those without independent mobility difficulties. Third, we performed gender-subgroup analyses for the association between driving status and visiting places.

Furthermore, we exploratory conducted moderation analyses by alternative transportation means (public transportation and ride-sharing with family/friends). In the same analytical model, we performed an analysis that included a product term of driving status and public transportation or ride-sharing with family/friends (code: not available=0 and available=1).

To mitigate the potential bias due to the missing information, the multiple imputation approach was applied under the Missing at Random (MAR) assumption. We generated 20 imputed datasets using the multiple imputation by chained equations (MICE) procedure and pooled the results by the standard Rubin’s rule [38].

The statistical significance was set at *P*<0.05. All statistical analyses were conducted using R software (Version 4.3.2 for Windows; R Foundation for Statistical Computing, Vienna, Austria).

## Results

A total of 432 individuals were analyzed. Table 1 presents the characteristics of the participants. The mean age (SD) was 74.8 (6.0) years, and 52.8% were women. Among the participants, 24.5% were non-drivers, and 67.1% for public transportation and 64.8% for car rides with family or friends were available. Non-drivers were more likely to be older, female, live alone, have lower incomes, have more illnesses, have declining mental health, and less impaired IADL performance. Additionally, non-drivers were more likely to be available for car rides with family/friends and public transportation.

**Table 1.**
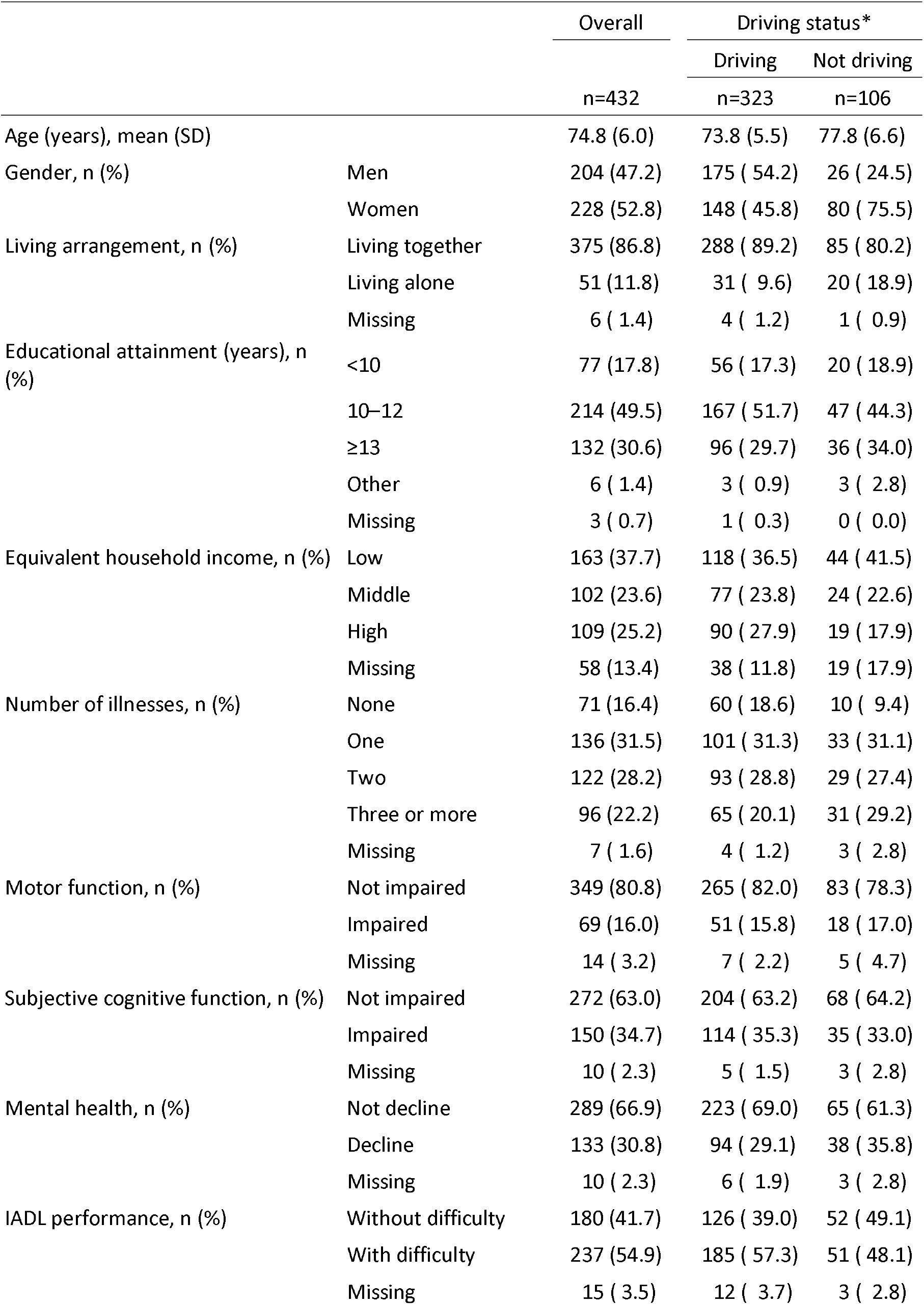
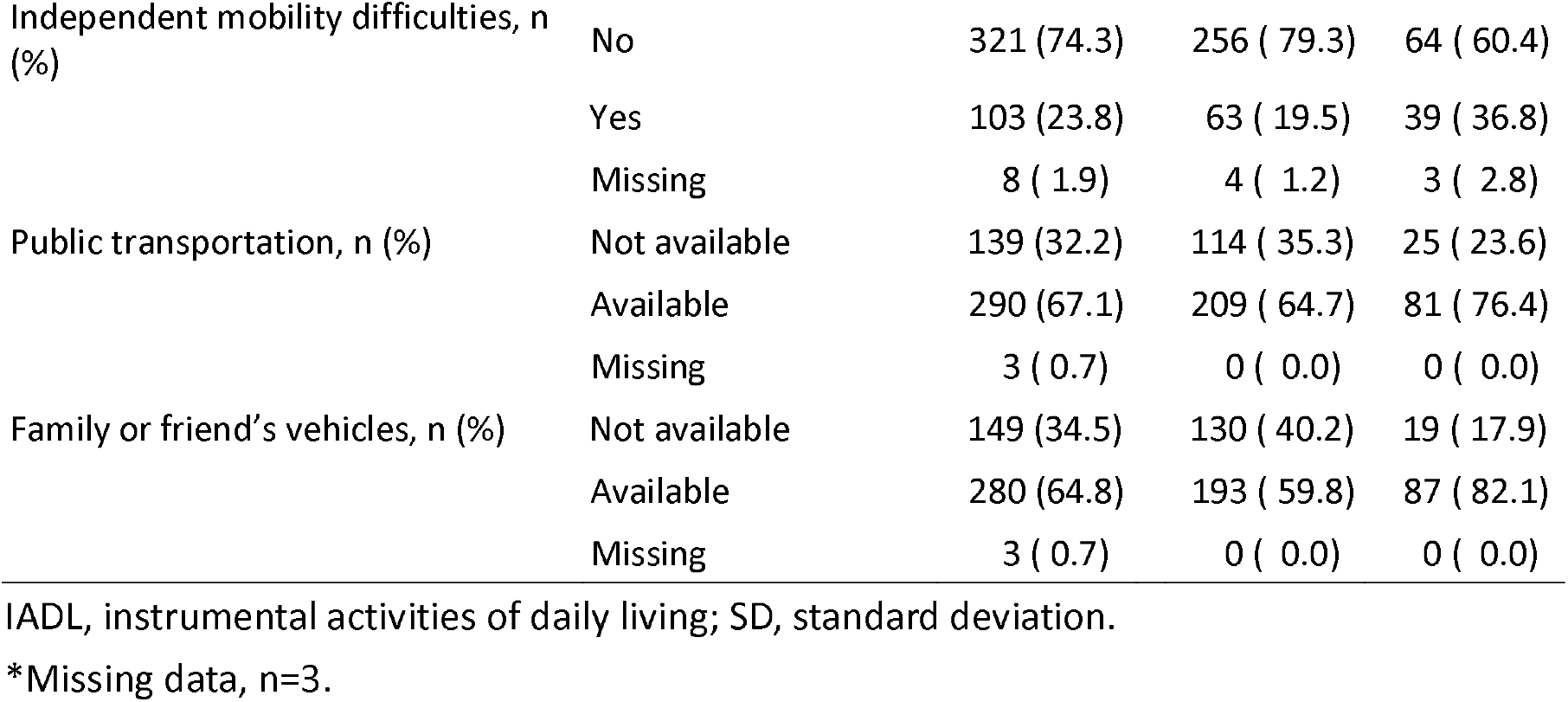
Characteristics of the participants.

Table 2 shows the descriptive statistics of the scores for visiting places (Supplementary Table 1 shows the results for all 34 places). The mean score (SD) of the overall participants was 21.5 (5.2). The mean score of drivers was 22.0 (4.8), while that of non-drivers was 20.0 (5.9). Regarding the domain of consumer, administration, and self-care places, overall participants’ mean score (SD) was 8.2 (1.3), drivers’ mean score was 8.3 (1.1), and non-drivers’ mean score was 7.9 (1.7). For the domain of places for medical and health care, overall participants’ mean score (SD) was 2.5 (0.9), drivers’ mean score was 2.5 (0.9), and non-drivers’ mean score was 2.5 (0.9). Regarding the domain of social, cultural, and spiritual places, overall participants’ mean score was 5.9 (2.4), drivers’ mean score was 6.1 (2.4), and non-drivers’ mean score was 5.2 (2.6). Finally, for the domain of places of recreation and physical activities, overall participants’ mean score (SD) was 4.8 (2.1), drivers’ mean score was 4.9 (2.1), and non-drivers’ mean score was 4.2 (2.0).

**Table 2.** Descriptive statistics of the scores of the visiting places.

|  | Overall | Driving status |  |
| --- | --- | --- | --- |
|  |  | Driving | Not driving |
|  | Mean (SD) | Mean (SD) | Mean (SD) |
| All places (0–34 points) | 21.5 (5.2) | 22.0 (4.8) | 20.0 (5.9) |
| Subdomains |  |  |  |
| Consumer, administration, and self-care places (0–9 points) | 8.2 (1.3) | 8.3 (1.1) | 7.9 (1.7) |
| Places for medical and health care (0–5 points) | 2.5 (0.9) | 2.5 (0.9) | 2.5 (0.9) |
| Social, cultural, and spiritual places (0–12 points) | 5.9 (2.4) | 6.1 (2.2) | 5.2 (2.6) |
| Places of recreation and physical activities (0–8 points) | 4.8 (2.1) | 4.9 (2.1) | 4.2 (2.0) |

SD, standard deviation.
Missing data: driving status (n=3), scores of the visiting places (all places, n=35; consumer, administration, and self-care places, n=11; places for medical & health care, n=12; social, cultural, spiritual places, n=13; places of recreation and physical activities, n=17).

Table 3 demonstrates the association between driving status and the scores for visiting places, based on multivariable linear regression analysis. After adjusting for all the covariates, the analysis revealed that not driving was significantly associated with lower scores for visiting places (β=-0.40; 95% CI=-0.61, -0.19; *P*<0.001). Regarding the subdomains, not driving was significantly associated with lower scores for consumer, administration, and self-care places (β=-0.32; 95% CI=-0.55, -0.09; *P*=0.007) and social, spiritual, and cultural places (β=-0.44; 95% CI=-0.66, -0.23; *P*<0.001). Regarding the other subdomains, not driving showed low visiting place scores, but their significant associations were not found. These results were replicated in the sensitivity analyses using IPW (Supplementary Table 2). Additionally, the analysis that excluded individuals with independent mobility difficulties yielded similar results (Supplementary Table 3). Furthermore, the gender-subgroup analysis demonstrated similar tendencies in both men and women, although the associations were slightly larger for men (Supplementary Table 4).

**Table 3.** Association between driving status and the visiting places, based on multivariable linear regression analysis.

|  | All places |  | Subdomains |  |  |  |  |  |  |  |
| --- | --- | --- | --- | --- | --- | --- | --- | --- | --- | --- |
|  |  |  | Consumer, administration,<br>and self-care places |  | Places for medical and<br>health care |  | Social, cultural, and spiritual<br>places |  | Places of recreational and<br>physical activities |  |
| | $\beta$ (95% CI) | *P*-value | $\beta$ (95% CI) | *P*-value | $\beta$ (95% CI) | *P*-value | $\beta$ (95% CI) | *P*-value | $\beta$ (95% CI) | *P*-value |
| Crude | -0.39 (-0.61, -0.17) | < 0.001 | -0.30 (-0.52, -0.08) | 0.007 | -0.03 (-0.26, 0.19) | 0.778 | -0.39 (-0.61, -0.17) | < 0.001 | -0.30 (-0.52, -0.07) | 0.009 |
| Adjusted | -0.40 (-0.61, -0.19) | < 0.001 | -0.32 (-0.55, -0.09) | 0.007 | -0.11 (-0.36, 0.14) | 0.390 | -0.44 (-0.66, -0.23) | < 0.001 | -0.23 (-0.46, 0.00) | 0.056 |

$\beta$ , regression coefficient; CI, confidence interval.
The visiting place scores were entered into the regression model after standardization.
Adjusted for age, gender, living arrangement, educational attainment, equivalent household income, comorbidities, instrumental activities of daily living performance, motor function, subjective cognitive function, mental health, independent mobility difficulties, public transportation availability, and families or friends' vehicle availability.
Missing data were imputed by multiple imputation approach.

Table 4 presents the results of moderation by alternative means of transportation for the association between driving status and visiting places. The availability of public transportation has moderated the decrease in the scores of visiting places for non-drivers (P for interaction=0.006). Additionally, it mitigated the decline in scores of the subdomains of the consumer, administration, and self-care places (P for interaction<0.001) and places for medical and health care (P for interaction=0.045). However, the availability of ride-sharing with family/friends did not moderate the reduction in the total and subdomain scores for visiting places as the participants were due to not driving..

**Table 4.** Moderation by other transportation means on the association between driving status and the visiting places, based on ltivariable linear regression analysis.

|  | All places |  | Subdomains |  |  |  |  |  |  |  |
| --- | --- | --- | --- | --- | --- | --- | --- | --- | --- | --- |
|  |  |  | Consumer, administration, and self-care places |  | Places for medical and health care |  | Social, cultural, and spiritual places |  | Places of recreational and physical activities |  |
|  | β (95% CI) | P-value | β (95% CI) | P-value | β (95% CI) | P-value | β (95% CI) | P-value | β (95% CI) | P-value |
| Public transportation |  |  |  |  |  |  |  |  |  |  |
| Available | -0.24 (-0.47, -0.01) | 0.043 | -0.05 (-0.25, 0.15) | 0.597 | -0.02 (-0.29, 0.25) | 0.885 | -0.29 (-0.55, -0.03) | 0.027 | -0.21 (-0.47, 0.05) | 0.119 |
| Not available | -0.68 (-1.16, -0.20) | 0.007 | -0.80 (-1.43, -0.18) | 0.014 | -0.42 (-1.02, 0.18) | 0.173 | -0.68 (-1.12, -0.25) | 0.003 | -0.20 (-0.74, 0.35) | 0.483 |
|  | P for interaction = 0.006 |  | P for interaction < 0.001 |  | P for interaction = 0.045 |  | P for interaction = 0.108 |  | P for interaction = 0.411 |  |
| Family or friend's vehicles |  |  |  |  |  |  |  |  |  |  |
| Available | -0.47 (-0.72, -0.22) | < 0.001 | -0.37 (-0.66, -0.09) | 0.010 | -0.15 (-0.43, 0.12) | 0.278 | -0.48 (-0.73, -0.22) | < 0.001 | -0.31 (-0.58, -0.04) | 0.026 |
| Not available | -0.36 (-0.79, 0.08) | 0.113 | -0.21 (-0.66, 0.23) | 0.352 | -0.11 (-0.69, 0.47) | 0.709 | -0.53 (-1.00, -0.07) | 0.026 | -0.08 (-0.57, 0.40) | 0.736 |
|  | P for interaction = 0.483 |  | P for interaction = 0.785 |  | P for interaction = 0.206 |  | P for interaction = 0.509 |  | P for interaction = 0.686 |  |

$\beta$ , regression coefficient; CI, confidence interval.
The visiting place scores were entered into the regression model after standardization.
Adjusted for age, gender, living arrangement, educational attainment, equivalent household income, comorbidities, instrumental activities of daily living performance, motor function, subjective cognitive function, mental health, independent mobility difficulties, and public transportation or families or friends' vehicle availability.
Missing data were imputed by multiple imputation approach.

## Discussion

This study examined the association between driving status and visiting places among older adults in a suburban area of Japan. Additionally, we explored the moderating role of alternative transportation means in this association. The results indicated that not driving was negatively associated with more visiting places, whereas public transportation availability, including trains and buses, moderated this adverse association. Our findings suggest that driving restrictions have the potential to reduce older adults’ places to visit, highlighting the necessity of developing a supportive environment that maintains and promotes their places of living and activities, regardless of mobility resources.

Driving is a major means of travel, particularly for older adults in rural and suburban areas [8, 9]. Several previous studies have shown that driving restrictions lead to decreasing social participation and outgoing frequencies among older adults [16, 17, 26]. Our results are in line with the evidence of decreased activity in older adults owing to limited driving, highlighting disparities in their visiting places depending on driving resources. Given that the places where individuals spend time in familiar spaces and engage in meaningful activities within the community can form an individual’s identity [18], shrinking their boundaries by limiting the places they visit may cause a crisis to their identity. With driving restrictions reducing the range of their living activities, the limited places of visit may compromise an individual’s meaning of life in their community.

In the present study, we also identified the types of visiting places affected by driving restrictions. The domain of consumer, administration, and self-care places includes essential living facilities and places, such as grocery stores, banks, and government offices. In particular, when shopping for food, the lack of a car to transport packages may increase the use of other means such as delivery, which might reduce the frequency of visits to these places. Alternatively, the negative effects of driving restrictions may depend on whether the facility or place is within walking distance. As the results for each place indicate (Supplementary Table 1), facilities that are less abundant in the area, such as government offices or malls, may be noticeably affected by restrictions on car mobility. Meanwhile, the domain of social, cultural, and spiritual places contains friends’ homes and leisure facilities and places. A study in the suburbs of the United Status shows that non-driving older adult are less likely to travel for leisure, which is in line with our results [22]. Considering that visits to these places are not necessarily essential in daily life, restrictions on flexible travel means may substantially affect visits to these places. This means that the demand for visits becomes latent due to driving restrictions. Nevertheless, although it does not interfere with their lives, restricting visits to such leisure-related sites may reduce the frequency of social interactions and meaningful activities, thereby decreasing their well-being in community life. These findings highlight the importance of carefully evaluating and supporting affected places to visit for older adults with limited driving.

This study did not support associations between driving status and the domains of medical and healthcare places and recreation and physical activity places. Regarding medical and health care, especially in old age, it may be necessary to visit the places regularly for medical examinations, doctor visits, and health checkups. A greater need for life might prompt the use of other means of transportation for visits, even if they are costly; explicit demand increases the priority of the visit. Thus, it is plausible that a noticeable effect of driving restrictions could not be observed in medical and healthcare places. For the domain of recreation and physical activity places, which includes green and blue areas and sports facilities, the analysis found a marginal association between not driving and fewer visits to these places. Although the present study could not confirm this, not driving might also have slightly limited visits to places related to recreation and physical activity.

In summary, our findings suggest that driving restrictions may reduce older adults’ visiting places, and particularly, indicate places of life-related and leisure and cultural places as susceptible domains. Policymakers in public health and urban planning should therefore understand the distinctive effects of driving restrictions on older adults outside the home and provide them with a physical and social environment to ensure mobility support for their well-being.

The present study found that public transportation availability (e.g., public trains and buses) moderated the association between not driving and fewer visiting places. The availability of public transport has a role to play in encouraging older adults to go outside and engage in activities [25]. Public transportation promotes healthcare access and social engagement in older adults, which may reduce isolation and improve mental health [39]. Although the benefits of public transportation do not necessarily apply to physical activities and health status [26-28], our findings suggest that public transportation can help non-drivers sustain places to visit. Because public transportation can have meaning in replacing non-driving older adults visiting various places, ensuring its accessibility and affordability is a priority for older adults, especially those who do not drive. Nevertheless, given that suburban and rural areas do not always have sufficient resources for public transport, their accessibility depends on the policy direction and resources of the area. Informal mobility resources, such as family/friends’ vehicles, are expected to be an alternative; however, our results showed that the availability of ride-sharing with family/friends did not moderate the adverse associations between not driving and visiting places. Although informal transport resources are a crucial means of mobility for older adults with driving restrictions, especially in non-urban areas, it has been pointed out that those who depend on informal means, such as ride-sharing with family/friends, may experience a latent demand for travel (i.e., giving up or canceling going out) [40]. The availability of informal means is highly dependent on social relationships; given their time and behavioral constraints, informal options might not universally complement the negative influence of driving restrictions on visiting places. Considering that the use of informal transportation keeps older adults active, informal resources may have some role in meeting the needs for basic outings. However, they may not adequately address the decrease in visiting places affected by driving restrictions. Still, we could not rule out the possibility that these results depended on the survey area of a suburban having a certain amount of public transportation resources. Suburban and rural areas with fewer public transport resources might have greater relative importance for alternative resources, such as informal options, thus requiring future investigations in more diverse areas. Even then, building age-friendly communities requires an understanding of the unmet needs of older adults and establishing alternatives to compensate for these needs, including government support and fostering informal mobility assistance.

There are several limitations in this study. First, the nature of the observational cross-sectional study design could not determine the causality. The reverse causation should be noted. Additionally, we did not have any information on past driving status or the restricted periods. Further studies using longitudinal data are thus required to determine the effect of driving limitations on visiting places. Second, the participants were recruited from individuals living in a suburban area. It is unclear whether the results can be applied to urban or rural areas, thus limiting their transportability. Third, this study did not assess detailed information about the places to visit (i.e., whether the place was meaningful or familiar to the participants, and what activities they performed there). Further research with a more detailed assessment is required to confirm this finding. Finally, this study was conducted in a single municipality in Japan, which restricts the generalizability of the results.

Despite the above limitations, this study holds significance in the first to identify the places that non-driving older adults visit less, emphasizing the importance of promoting an accessible environment regardless of driving ability. Especially, social, spiritual, and cultural places, although not necessarily essential for daily life, are likely to be places with social interactions and meaningful activities that are crucial to the well-being of older adults. Understanding the importance of these non-essential places can encourage efforts to improve access to these places. Meanwhile, public transportation can be meaningful in supporting visiting places. Promoting age-friendly cities emphasizes meeting the unmet transport needs of older adults, which requires the provision of a range of mobility alternatives, including existing public transportation; for instance, some municipalities in Japan are operating new transportation systems, such as on-demand buses and green slow mobility [4]. It is necessary to provide diverse and affordable means of transportation to ensure that older adults can visit various places.

In conclusion, this study revealed that not driving was negatively associated with higher visiting places among older adults in a suburban area. Particularly, non-driving individuals were less likely to visit places related to consumer, administration, and self-care, as well as places related to social, spiritual, and cultural activities. Meanwhile, the availability of public transport moderated the associations between not driving and fewer visiting places. These findings suggest the importance of developing physical and social environments, including other mobility resources, to support older adults with driving limitations in going out and visiting places.

## Data Availability

The data supporting the findings of this study are not publicly available because of privacy or ethical restrictions.

## Statements

## Acknowledgement

We wish to express our sincere gratitude to Dr. Lee Sangyoon, Center for Dementia Care Research and Practices and National Center for Geriatrics and Gerontology, for contribution to this study. We also thank the staff of the Obu City Government Office for supporting this study and all the participations.

## Statement of Ethics

Study approval statement: This study was reviewed and approved by the Research Ethics Committee of the National Centre for Geriatrics and Gerontology (approval no. 1774). The study was conducted in accordance with the principles of the Declaration of Helsinki.

Consent to participate statement: Written informed consent was obtained from all participants.

## Author Contributions

TN (Taiji Noguchi) conceptualized and designed the study, analyzed the data, interpreted the results, and drafted and revised the manuscript. AK conceptualized and designed the study, participated the data collection, supported the analysis of the data, interpreted the results, and drafted and revised the manuscript. SO conceptualized the study, participated in data collection, supported the interpretation of the results, and reviewed and critically revised the manuscript. XJ, TN (Takeshi Nakagawa), and YS supported data collection and reviewed and critically revised the manuscript. TS, the principal investigator of the NCGG-UniCo project, conceptualized and designed the study, participated in the data collection, supported data analysis, interpreted the results, and drafted and revised the manuscript.

