## supplementary for "Association between driving status and visiting places among older adults in a suburban area in Japan: Findings from a cross-sectional survey"

**Supplementary Table 1. Prevalence of the visiting places (%)**

|  |  |  | Driving status | | |
| --- | --- | --- | --- | --- | --- |
|  | Overall |  | Driving |  | Not driving |
| **Consumer, administration, and self-care places** | | | |  |  |
| Small grocery shop | 96.5 |  | 96.6 |  | 96.2 |
| Small shop | 92.7 |  | 93.4 |  | 90.5 |
| Supermarket | 97.7 |  | 98.5 |  | 95.2 |
| Convenience store | 95.3 |  | 96.6 |  | 91.4 |
| Mall | 82.7 |  | 84.2 |  | 77.9 |
| Pharmacy | 95.8 |  | 96.9 |  | 93.3 |
| Hairdresser, salon, barbershop | 86.3 |  | 86.0 |  | 87.7 |
| Bank, post office | 94.7 |  | 95.7 |  | 91.5 |
| Government office | 81.6 |  | 85.7 |  | 68.6 |
| **Places for medical care and health care** | |  |  |  |  |
| Alternative adjuvant therapy | 23.4 |  | 24.1 |  | 21.4 |
| Dentist's office | 78.7 |  | 78.6 |  | 78.8 |
| Doctor's office | 91.2 |  | 91.0 |  | 91.5 |
| Hospital, health center | 58.7 |  | 59.4 |  | 55.9 |
| Senior day care | 0.9 |  | 0.3 |  | 2.9 |
| **Social, cultural, and spiritual places** | |  |  |  |  |
| Family/relative's place | 86.4 |  | 88.8 |  | 79.6 |
| Friend's place | 70.4 |  | 73.4 |  | 61.5 |
| Restaurant, café | 93.0 |  | 95.0 |  | 86.7 |
| Bar | 18.7 |  | 22.6 |  | 6.8 |
| Karaoke box | 8.6 |  | 9.3 |  | 6.8 |
| Community center | 63.0 |  | 62.2 |  | 65.7 |
| Building for worship, cemetery | 86.5 |  | 88.9 |  | 79.0 |
| Library | 33.2 |  | 32.8 |  | 34.0 |
| Entertainment, cultural places | 48.0 |  | 50.8 |  | 40.4 |
| Historic building | 39.1 |  | 41.9 |  | 31.1 |
| Stadium, theater | 33.6 |  | 37.6 |  | 20.6 |
| Pinball game place | 7.5 |  | 9.3 |  | 2.0 |
| **Places for recreation and physical activities** | | | |  |  |
| Neighborhood | 91.5 |  | 92.2 |  | 89.3 |
| Garden | 45.6 |  | 48.8 |  | 35.9 |
| Park, green area | 82.3 |  | 82.3 |  | 81.9 |
| Sports facility | 35.6 |  | 36.6 |  | 31.1 |
| Hot spring, public bath | 45.7 |  | 49.8 |  | 32.7 |
| Cottage, summer house | 47.7 |  | 51.7 |  | 35.0 |
| Forest, mountain, lake, sea | 56.4 |  | 60.2 |  | 43.7 |
| Transportation center | 71.7 |  | 70.4 |  | 75.0 |

**Supplementary Table 2. Association between driving status and the visiting places, based on linear regression analysis with inverse probability weighting**

| All places | |  | Subdomains | | | | | | | | | | |
| --- | --- | --- | --- | --- | --- | --- | --- | --- | --- | --- | --- | --- | --- |
|  | |  | Consumer, administration, and self-care places | |  | Places for medical and health care | |  | Social, cultural, and spiritual places | |  | Places of recreational and physical activities | |
| β (95% CI) | *P*-value |  | β (95% CI) | *P*-value |  | β (95% CI) | *P*-value |  | β (95% CI) | *P*-value |  | β (95% CI) | *P*-value |
| -0.27 (-0.48, -0.06) | 0.013 |  | -0.26 (-0.46, -0.06) | 0.011 |  | 0.01 (-0.21, 0.22) | 0.947 |  | -0.30 (-0.52, -0.09) | 0.006 |  | -0.17 (-0.39, 0.04) | 0.116 |

β, regression coefficient; CI, confidence interval.

The visiting place scores were entered into the regression model after standardization.

The regression model was performed with stable inverse probability weighting to balance the background factors for each driving status after multiple imputation approach.

**Supplementary Table 3. Restricted analysis of those without independent mobility difficulties for the association between driving status and the visiting places, based on multivariable linear regression analysis**

|  | All places | |  | Subdomains | | | | | | | | | | |
| --- | --- | --- | --- | --- | --- | --- | --- | --- | --- | --- | --- | --- | --- | --- |
|  |  | |  | Consumer, administration, and self-care places | |  | Places for medical and health care | |  | Social, cultural, and spiritual places | |  | Places of recreational and physical activities | |
|  | β (95% CI) | *P*-value |  | β (95% CI) | *P*-value |  | β (95% CI) | *P*-value |  | β (95% CI) | *P*-value |  | β (95% CI) | *P*-value |
| Adjusted | -0.37 (-0.60, -0.14) | 0.002 |  | -0.28 (-0.49, -0.07) | 0.009 |  | -0.12 (-0.42, 0.17) | 0.417 |  | -0.37 (-0.62, -0.12) | 0.004 |  | -0.26 (-0.54, 0.02) | 0.070 |

β, regression coefficient; CI, confidence interval.

The visiting place scores were entered into the regression model after standardization.

Adjusted for age, gender, living arrangement, educational attainment, equivalent household income, comorbidities, instrumental activities of daily living performance, motor function, subjective cognitive function, mental health, public transportation availability, and families or friends’ vehicle availability.

Missing data were imputed by multiple imputation approach.

**Supplementary Table 4. Gender-subgroup analysis of the association between driving status and the visiting places, based on multivariable linear regression analysis**

|  | All places | |  | Subdomains | | | | | | | | | | |
| --- | --- | --- | --- | --- | --- | --- | --- | --- | --- | --- | --- | --- | --- | --- |
|  |  | |  | Consumer, administration, and self-care places | |  | Places for medical and health care | |  | Social, cultural, and spiritual places | |  | Places of recreational and physical activities | |
|  | β (95% CI) | *P*-value |  | β (95% CI) | *P*-value |  | β (95% CI) | *P*-value |  | β (95% CI) | *P*-value |  | β (95% CI) | *P*-value |
| Men | -0.58 (-0.98, -0.17) | 0.006 |  | -0.57 (-1.04, -0.1) | 0.018 |  | -0.35 (-0.78, 0.08) | 0.114 |  | -0.47 (-0.91, -0.04) | 0.035 |  | -0.36 (-0.76, 0.05) | 0.087 |
| Women | -0.33 (-0.57, -0.08) | 0.010 |  | -0.23 (-0.47, 0.01) | 0.057 |  | 0.01 (-0.31, 0.34) | 0.934 |  | -0.41 (-0.67, -0.16) | 0.002 |  | -0.19 (-0.48, 0.1) | 0.207 |
|  | *P* for interaction = 0.179 | |  | *P* for interaction = 0.178 | |  | *P* for interaction = 0.154 | |  | *P* for interaction = 0.713 | |  | *P* for interaction = 0.287 | |

β, regression coefficient; CI, confidence interval.

The visiting place scores were entered into the regression model after standardization.

Adjusted for age, living arrangement, educational attainment, equivalent household income, comorbidities, instrumental activities of daily living performance, motor function, subjective cognitive function, mental health, independent mobility difficulties, public transportation availability, and families or friends’ vehicle availability.

Missing data were imputed by multiple imputation approach.
